# GPT for RCTs?: Using AI to determine adherence to reporting guidelines

**DOI:** 10.1101/2023.12.14.23299971

**Authors:** J.G. Wrightson, P. Blazey, D. Moher, K.M. Khan, C.L. Ardern

## Abstract

**Background:** Adherence to established reporting guidelines can improve clinical trial reporting standards, but attempts to improve adherence have produced mixed results. This exploratory study aimed to determine how accurate a Large Language Model generative AI system (AI-LLM) was for determining reporting guideline compliance in a sample of sports medicine clinical trial reports.

**Design and Methods:** This study was an exploratory retrospective data analysis. The OpenAI GPT-4 and Meta LLama2 AI-LLMa were evaluated for their ability to determine reporting guideline adherence in a sample of 113 published sports medicine and exercise science clinical trial reports. For each paper, the GPT-4-Turbo and Llama 2 70B models were prompted to answer a series of nine reporting guideline questions about the text of the article. The GPT-4-Vision model was prompted to answer two additional reporting guideline questions about the participant flow diagram in a subset of articles. The dataset was randomly split (80/20) into a TRAIN and TEST dataset. Hyperparameter and fine-tuning were performed using the TRAIN dataset. The Llama2 model was fine-tuned using the data from the GPT-4-Turbo analysis of the TRAIN dataset. Primary outcome measure: Model performance (F1-score, classification accuracy) was assessed using the TEST dataset.

**Results:** Across all questions about the article text, the GPT-4-Turbo AI-LLM demonstrated acceptable performance (F1-score = 0.89, accuracy[95% CI] = 90%[85-94%]). Accuracy for all reporting guidelines was > 80%. The Llama2 model accuracy was initially poor (F1-score = 0.63, accuracy[95%CI] = 64%[57-71%]), and improved with fine-tuning (F1-score = 0.84, accuracy[95%CI] = 83%[77-88%]). The GPT-4-Vision model accurately identified all participant flow diagrams (accuracy[95% CI] = 100%[89-100%]) but was less accurate at identifying when details were missing from the flow diagram (accuracy[95% CI] = 57%[39-73%]).

**Conclusions:** Both the GPT-4 and fine-tuned Llama2 AI-LLMs showed promise as tools for assessing reporting guideline compliance. Next steps should include developing an efficent, open-source AI-LLM and exploring methods to improve model accuracy.

## Introduction

Poor reporting of clinical trials is common [1], threatens the reliability and credibility of medical research [2] and affects patient care [3]. Improving trial reporting, therefore, is an ethical imperative [4,5]. Using reporting guidelines, such as the CONsolidated Standards of Reporting Trials (CONSORT), can improve trial reporting standards [6–8], but adherence is often poor [9]. Following recent calls to evaluate the role of Artificial Intelligence (AI) in facilitating editorial and peer review decisions [10], we assessed how well an AI model could determine reporting guideline adherence in clinical trial reports.

Clinical trial reporting standards are improved when medical journals instruct authors to use reporting guideline checklists, but not all journals require adherence to reporting guidelines, and enforcement of adherence is inconsistent [11–14]. Recent attempts to improve reporting standards involve training peer reviewers, authors, or editors, in the use of reporting guideline checklists with mixed results [15,16]. It might be possible to improve guideline adherence by providing feedback to authors, either prior to submission or during peer review [16].

Using machine learning and AI, especially Large Language Model generative AI systems (AI-LLM), to check for adherence to reporting guidleines might save authors, reviewers and publishers time and make the editorial process more efficient [17,18]. An AI-LLM can discern— with 80-90% accuracy—whether the content of computer science manuscripts corresponds to reporting guideline checklists [17]. Given this success, generative AI systems hold promise for evaluating and improving adherence to reporting guidelines.

There is increasing interest in the rigour and transparency, or lack thereof, of sports medicine, exercise science and orthopedic research [19]. Reporting in sports medicine and exercise science papers is often inadequate, and there are concerns about the reproducibility and veracity of many findings [19–23]. Improving reporting practices should be a priority for researchers and publishers [19,24] in all fields, including sports medicine. This exploratory research aimed to answer the following research question: How accurately can an AI-LLM measure reporting guideline compliance in a sample of sports medicine clinical trial reports?

## Method

This study was an exploratory retrospective data analysis. The study is reported in accordance with the Minimum Information about CLinical Artificial Intelligence Modeling (MI-CLAIM) standards [25], and a completed MI-CLAIM checklist is available on the Open Science Framework (OSF; https://tinyurl.com/gpt4rct1).

### Pilot study

In a pilot study, we tested whether an earlier version of OpenAI’s GPT AI-LLM (GPT 3.5, San Francisco, USA) could measure reporting guideline compliance in a sample of Sports Medicine clinical trial reports. Details of the methods and results of that study can be found on the OSF. We used the same data in the present study.

### Data

We used a sub-sample of the dataset provided by Schulz et al. (2022) [19]. In their systematic review, Schulz and colleagues analyzed the reporting practices, including items from the CONSORT checklist, of 160 peer-reviewed scientific papers published in Sports Medicine journals in 2020. Journals were identified using the Scimago Journal Rank indicator (see Schulz et al. 2022 [19] for details). We extracted all papers from the Schulz et al. dataset that were available in full-text machine-readable format. Details for the data extraction are shown in the R notebooks located on the OSF: https://tinyurl.com/gpt4rctdata.

The data for open-access papers (n=24) were extracted from the PubMed Central database in machine-readable form. The data for the remaining papers were extracted from articles with electronic (‘Epub’) or PDF files accessible by the study lead author (JW). Papers were removed from analysis if a) the text extraction contained errors or b) the electronic file was inaccessible. The by-journal distribution of papers included in the analysis is shown in Figure 1. Data were split into TRAIN (80% of text-question pairs) and TEST (20%) datasets. The split was stratified across the paper sections (Introduction/Method/Results) so that a single paper could be used in both the TEST and TRAIN datasets (e.g., the Method section in the TEST dataset and the Results section in the TRAIN dataset). The characteristics of the datasets are shown in the Supplementary Materials Table S1. We did not create a validation data set because of the relatively low number of training examples.

**Figure 1.**
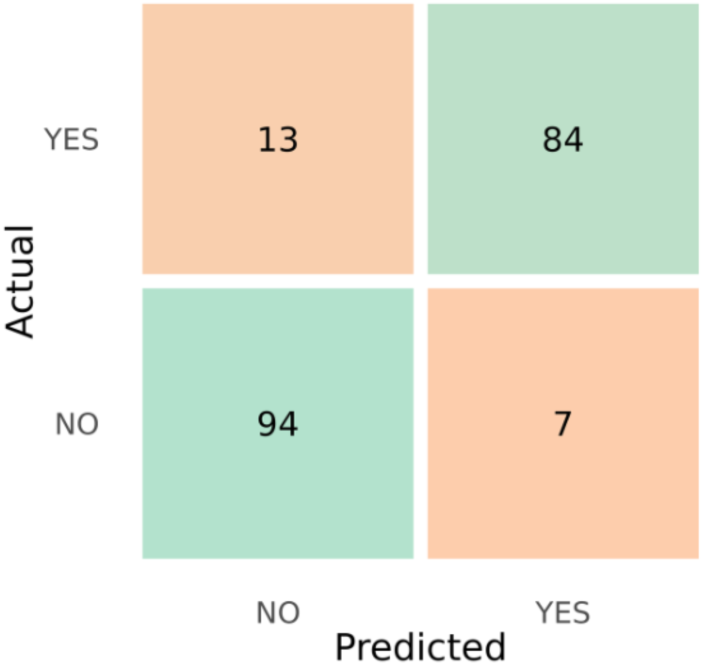
Confusion matrix from the analysis of the TEST dataset. Actual (y-axis) represents the answers given by the GPT-4-turbo model, and Predicted (x-axis) represents the labels for each question extracted from Schulz et al (2022).

Data extraction

The limit on the size of data (the ‘context window’) submitted to the GPT 3.5 (from the pilot study) and Meta’s Llama 2 (Menlo Park, USA) AI models meant that entire papers could not be analyzed. Instead, we followed the example of Liu and Shah [17] and divided each paper into three sections: the Introduction, Method and Results. For each paper, nine pairs of a section of text from the paper (e.g. methods) and a question about the text (text-question pairs) were created to match the reporting guideline items that could be assessed using the AI model (see Table 1 below). For one item (see Question 9, Table 2) we combined the method and results section of the paper. This was necessary because the label provided in the Schulz et al (2022) data specifically referred to the primary outcome, which may only have been identified in the method section of each paper. This question could not be analyzed using the Llama 2 model because of the smaller context window allowed for this model. The model response was limited to 512 tokens. Only the text- question pair was removed at this stage, and thus, in the final analysis, some papers did not have all the text-question pairs (Supplementary Material Table S1).

**Table 1.** The number of text-question pairs for each reporting guideline in the dataset, and the proportion of papers that adhered to each guideline.

| # | Paper Section | Reporting Guideline Item | Pairs (n) | Adherence (%) |
| --- | --- | --- | --- | --- |
| 1 | Introduction | The study hypotheses | 113 | 65% |
| 2 | Method | The primary outcome | 108 | 50% |
| 3 | Method | The sample size | 108 | 67% |
| 4 | Method | The eligibility criteria | 108 | 77% |
| 5 | Method | The implementation of the randomization sequence | 108 | 58% |
| 6 | Method | The randomization methods | 108 | 41% |
| 7 | Method | The random allocation sequence & assignment | 108 | 26% |
| 8 | Method | Blinding | 108 | 49% |
| 9 | Results | Standardized effect sizes and confidence intervals | 113 | 20% |
| 10 | Results | The number of participants randomly assigned | 20* | 100%* |
| 11 | Results | The number of participants lost to follow-up | 20* | 50%* |

\* Subset of the full dataset, see 'Image analysis' for details

**Table 2.** Accuracy of the GPT-4 models for each reporting guideline item.

| # | Question | Accuracy [95%CI] |
| --- | --- | --- |
| **<u>Text Analysis</u>** |  |  |
| 1 | Does the article include the study hypotheses? | 83% [63-93%] |
| 2 | Does the article define the primary outcome or primary endpoint? | 89% [73-96%] |
| 3 | Does the Method include how the sample size was determined? | 90% [71-97%] |
| 4 | Does the article define the study's eligibility criteria? | 86% [67-95%] |
| 5 | Does the article provide a detailed description of the specific techniques used for the practical implementation of the randomization sequence? | 94% [72-99%] |
| 6 | Does the article include the randomization methods used in the study? | 94% [73-99%] |
| 7 | Does the article EXPLICITLY detail the individuals or bodies or roles involved in either GENERATING the random allocation sequence, or ENROLLING participants, or ASSIGNING participants to trial groups? | 86% [67-95%] |
| 8 | Does the article include details about who, if anyone, was blinded to the assignment to interventions? | 100% [87-100%] |
| 9 | For the PRIMARY outcome, do the results reported in the article include the standardized effect size and confidence interval? | 87% [68-95%] |
| **<u>Image Analysis</u>** |  |  |
| 10 | Does this image contain a CONSORT flow diagram showing, for each group, the number of participants who were randomly assigned, received the intended treatment, and were analysed for the primary outcome? | 100% [89-100%]* |
| 11 | Does this image display a CONSORT flow diagram that details both the number of participants lost to follow-up and excluded after randomization AND specifies the reasons for these losses in each group? | 57% [39-73%]* |

\* = Analysis performed on a subset (n=20) of the dataset

### Reporting Guideline Items and Data Labelling

The extracted papers were assessed for adherence to eleven reporting guideline items. These items included the nine ‘core’ Method and Results items from the CONSORT 2010 guidelines [16], with an additional item from the Introduction section (Table 1, Question 1) and Method section (Table 1, Question 3) sections. The text of each paper was assessed for adherence to nine reporting guideline items (Table 1, Items 1-9). The participant flow figures from a subset of the dataset were assessed for adherence to two further reporting items (Table 1, Items 10-11, see ‘Image Analysis’ below for details). Each reporting guideline item was assessed using a generative question and answering format, wherein the model was prompted to answer a question, formulated using natural language, about the text/image extracted from each paper. The model was required to summarize the text that was relevant to the question, and answer YES or NO to the question. Each question corresponded to a variable from the labelled dataset provided by Schulz et al (2022). The label (“ground truth”) for each question ("YES” or “NO”) was extracted from the systematic analysis by Schulz et al. (2022). Details for how each label was transformed into a “YES” or “NO” answer can be found in the raw data spreadsheets (https://tinyurl.com/gpt4rctdata).

### Model Choice and Optimization

We tested three models; OpenAI’s GPT-4-Turbo and GPT-4-Vision, and Meta’s Llama 2 70B. In our pilot study, we found that an earlier version of OpenAIs AI-LLM (GPT-3.5) could achieve adequate performance in the same task, but required significant tuning to do so. The newer GPT- 4 models perform significantly better on similar tasks [17,26]. We also tested the open-source Llama 2 AI-LLM. We included an open-source model because we envisage use cases where authors may want to run models locally or wish to support open-source systems in accordance with open science principles [27].

For the GPT-4 analysis, we used the hyperparameter settings from our pilot study (Temperature = 0.2 and Top-P = 0.2, see the pilot study https://tinyurl.com/gpt4rct1 for details of the hyperparameter tuning steps). Unfortunately, at the time of this analysis, we did not have access to fine-tune the GPT-4 model. Model tuning in the present study was achieved through iterative “prompt engineering”. We initially used the system and user prompts from our pilot study which were developed using the guidelines provided by OpenAI [28] and included asking the model to adopt a persona (the system prompt), using delimiters to distinguish parts of the input and specifying the steps required to complete the task (the user prompt). We then analyzed the training dataset using these prompts. We subsequently took the first 10 examples from the training dataset that the model had incorrectly answered, and used the OpenAI ChatGPT application to help us improve the system and user prompts by asking it to improve the language of the prompts to increase the likelihood that the model would respond correctly. The training dataset was rerun with these adjustments, and then a second round of examples was used to improve the prompts further. We used the text provided by ChatGPT for each prompt verbatim. The final system prompt attached to each question pair was as follows:

> *"You are a health researcher reviewing a scientific article for a peer-reviewed sports medicine journal. You will be supplied with text from the article, a description of a task, and a question (delimited by XML tags). Use only the information provided directly in the text to answer the question. Your answer must be accurate and precise. You must answer each question in steps. Delimit each step. Step 1: Summarize the information in the text that is relevant to the task description. Step 2: Answer the question with either ’YES’ or ’NO’ only."*

ChatGPT suggested providing a task description and a question in the user prompt. For example, the user prompt given to the model with the introduction text was formatted thus:

> *“Task: Analyze the article focusing specifically on the details of the hypotheses used in the study. Note that acceptable hypotheses are clear, testable propositions or predictions that are specifically formulated for statistical analysis. Your task is to identify whether the hypotheses are clearly stated and if they meet these criteria. If the text only mentions hypotheses in general without clearly stating them or if they are not formulated for statistical analysis, your answer should be ’NO’. Question: Does the article include the study hypotheses?"*

For clarity, only the questions included in the user prompt are shown in Table 1. The final full user prompt for each reporting guideline item is shown in the Supplementary Material Table S2.

The Llama 2 model was fine-tuned using the correctly answered questions from the GPT-4 analysis of the text training data. One of the reporting guideline items could not be included in this analysis (Table 2, Question 9) because this question required the text from the method and results to be passed to the model, exceeding the number of tokens allowed in the context window. The tuning was performed, and the model was hosted, on the TogetherAI platform (San Francisco, USA). The analysis of the TEST dataset using the base and fine-tuned Llama 2 models was performed using the TogtherAI playground using the platform default hyperparameter values (Temperature and Top-P = 0.7).

### Image analysis

During the study, OpenAI made their GPT-4-Vision model available for general use. We used this model to assess adherence to two further research guideline items (Table 2, Questions 10-11) related to the participant flow diagrams in a subset of papers (n = 20). We encoded the papers’ participant flow diagram images in base64 and appended the system prompt and item questions to the encoded image using the same ‘chat’ format API call used for the analysis of the text. Ten of the papers contained a flow diagram which showed the number of participants randomized into each group (answer “YES” to Table 2, Question 10) but did not include reasons for exclusions and loss to follow-up (answer “NO” to Table 2, Question 11). These papers were identified using the labels from Schulz et al (2022). Ten of these papers were in the dataset. The remainder had complete flow diagrams (answer “Yes” to both questions).

The PubMed Identifiers (PMID) for the papers included in the image analysis are available in the online materials. Because the number of suitable papers was so low, the included papers were drawn from both the test and training datasets. To ensure the model did not simply label any flow diagram as a participant flow diagram, a sample of 10 flow diagrams which did not show participant flow through a clinical trial was included in the dataset for one question (Table 2, Question 10). The details of these images are in the online materials.

### Analysis

Data extraction and analysis were performed using the R (version 4.3.2) and Python (version 3.8.17) programming languages. The primary outcome of this study was the F1-score (%) from the GPT-4 text analysis. The F1-score is the harmonic mean of the model precision (the ratio of true positives to the total number of identified positives) and recall (the ratio of true positives to the actual number of positive cases in the data). The F1-score controls for the expected class imbalances (YES or NO answers) in the dataset. The model classification accuracy (the ratio of true positives to the total number of cases) and associated 95% confidence interval (95%CI, [29]) were also calculated.

## Results

The breakdown of included papers (n = 113) by publication name is shown in the Supplementary Material Figure S1.

### Text analysis

Model performance for the text analysis was evaluated in the TEST (20% held back) dataset.

### GPT-4 Performance

The confusion matrix for the model performance in the analysis of the text from the TEST dataset is shown in Figure 1. The model performance on each question is shown in Table 2 (Questions 1-9) and in the Supplementary Material Table S3. Across all questions about the article text, the GPT-4-Turbo AI-LLM demonstrated acceptable performance (F1-score = 0.89, accuracy[95% CI] = 90%[85-94%]). Accuracy for the reporting guideline items ranged from 83-100%.

### Llama 2 performance

Llama 2 model performance was evaluated in eight questions. Model accuracy for each question is shown in the Supplementary Material Table S4. The accuracy of the base model was low (F1-score = 0.63, accuracy[95%CI] = 64%[57-71%]). Fine tuning the model improved accuracy (F1-score = 0.84, accuracy[95%CI] = 83%[77-88%]).

### Image analysis

The flow diagrams from a subset of the data (n = 20) were assessed for adherence to two further reporting guideline items (Questions 10-11, Table 2). The GPT-4-Vision model accurately identified all flowcharts (accuracy[95% CI] = 100%[89-100%]) but was inaccurate at identifying when details were missing from the flow chart (accuracy[95% CI] = 57%[39-73%]).

## Discussion

We wanted to determine how accurate AI-LLMs were for measuring reporting guideline compliance in a sample of clinical trial reports. The OpenAI GPT-4-Turbo AI-LLM achieved ∼90% accuracy across nine reporting guideline questions. Using an AI-LLM may help journal editors, publishers, peer reviewers and authors check reporting guideline adherence quickly and accurately, which could reduce both editorial workloads and waste in research[30].

Poor reporting of clinical trials negatively impacts downstream evidence synthesis, such as systematic reviews and clinical practice guidelines and can impact care and public trust in science [3,4,31,32]. Interventions to improve clinical trial reporting guideline adherence have been examined, but the results have been inconsistent [15]. Typically, these interventions target authors’ or peer reviewers’ behaviour [15,16] but may fail because they increase the already high workload for these groups [15]. Interventions targeting publishers are less common [16]. There are many automated tools that journal publishers and editors use to screen manuscripts for errors and misconduct [33]. Using a trained AI-LLM, publishers could screen submitted publications for adherence to relevant reporting guidelines and flag to authors, peer reviewers and editors where details may be missing. This approach could allow publishers to improve reporting standards without substantially increasing the workload on editors and peer reviewers.

The accuracy of the GPT-4 AI-LLM was slightly higher than the previous version of the model (GPT-3.5 ∼86%) from our pilot study (https://tinyurl.com/gpt4rct1) and that reported by Liu and Shah [17]. As expected, the Open-AI model was more accurate than the open-source Llama 2 model. However, fine-tuning the Llama 2 model significantly improved its accuracy. It is likely that any eventual tool that uses an AI-LLM to screen unpublished papers will be one in which the confidentiality of the text, and the geolocation where the application is hosted, can be controlled. For example, entities that publish clinical trials testing drug efficacy may not wish, or be allowed, to submit potentially sensitive information to corporations whose servers are across international borders. Open-source models will allow these stakeholders to host and train models in secure environments.

The development of models that allow users to pose natural language-style instructions and questions about images should expand the possibilities for AI-LLMs to evaluate the contents of scientific papers. We tested the model’s ability to evaluate participant flow diagrams, but these technologies could also be used to examine the figures provided as evidence of risk of bias in systematic reviews or the contents of forest plots in meta-analyses. In a small subset of data, we found that the GPT-4-vision model could identify all of the participant flow diagrams correctly. It was substantially less accurate when extracting information from the flow diagrams. This suggests that either the model will require tuning for it to be used to evaluate the contents of scientific figures and diagrams, or that at present it is not sophisticated enough to perform this task.

The less-than-perfect accuracy of the AI-LLMs and the tendency of the current generation of AI- LLMs to “hallucinate” content [34] means that human confirmation of compliance is required. However, academics and scientists have long used imperfect automated tools to assess reporting standards (e.g. [35]). Our results suggest a similar role for AI-LLMs in efforts to improve clinical trial reporting. To help more clearly define the efficacy and limitations of AI-LLMs, future research comparing custom, well-trained AI models with author-completed checklists is warranted.

The primary limitation of our study was the (small and homogenous) dataset. The homogenous nature of the clinical trials included in this dataset, and the poor adherence to reporting standards in sports medicine clinical trial reports [19,21,36] limits the generalizability of our findings to other fields. Indeed, it is likely that any future AI-LLM developed to assess reporting guideline adherence will need training on (much larger) datasets that include clinical trials from a broad range of disciplines. Given that at least 75 clinical trials are published daily, this task appears achievable.

It was beyond the scope of this exploratory study to create a large, well-labelled dataset for model tuning. The model was therefore trained using data generated by the model (the correct answers and extracted text from the training data), possibly increasing tendencies to hallucinate and other errors. Nevertheless, acceptable model performance was achieved, and results should improve with larger samples. We did not analyze full papers, something that is possible with GPT-4-Turbo because of its larger context window but not for the version of Llama 2 used here (though large context window Llama 2 models have been developed (e.g. https://tinyurl.com/gpt4rct3), though at the time of analysis we were unable to fine-tune these models on the TogetherAI platform.

It is likely that analysis of whole papers will be necessary if AI-LLM are to be used to evaluate reporting guidelines compliance. For example, it will be necessary to know what the study hypothesis was when evaluating the statistical methods, results and discussion. Beyond establishing quantitative information regarding the accuracy of using AI models to assist in an efficient assessment of the completeness of reporting clinical trials, it is also likely important to ask a broad spectrum of editors about accuracy thresholds they regard as minimal. For example, would there be consensus that 70% of a clinical trial report needs to adhere to a reporting guideline checklist, or should the consensus be 85%, before a journal agrees to consider the paper for peer review? When journals establish this information, prospective authors may wish to use the same tools to ensure their clinical report meets these thresholds.

## Conclusion

AI-LLMs were sufficiently accurate for assessing reporting guideline compliance in clinical trial reports. However, variations in accuracy across different items indicate that, at present, these tools can assist with, but not replace, human evaluation of reporting standards compliance.

## Data Availability

Code notebooks will be available on the Open Science Framework (https://tinyurl.com/gpt4rctdata) upon publication. Copyright issues prevent us from sharing the text extracted from the papers used in this analysis; however, details of the steps needed to reproduce the extracted text from open and closed papers can be found within these notebooks.

## Acknowledgements

All authors have seen and approved the manuscript form. We would like to thank Dr. Nihar Shah for his advice and feedback, and the organizers and attendees of the Meta-Research Innovation Center at Stanford University’s international forum for their comments and discussion of our work.

## Data availability

Code notebooks and data are available on the Open Science Framework (https://tinyurl.com/gpt4rctdata). Copyright issues prevent the sharing of some of the text extracted from the papers used in this analysis; however, details of the steps needed to reproduce the extracted text from open and closed papers can be found within these notebooks.

## Supplementary Material

**Figure S1.**
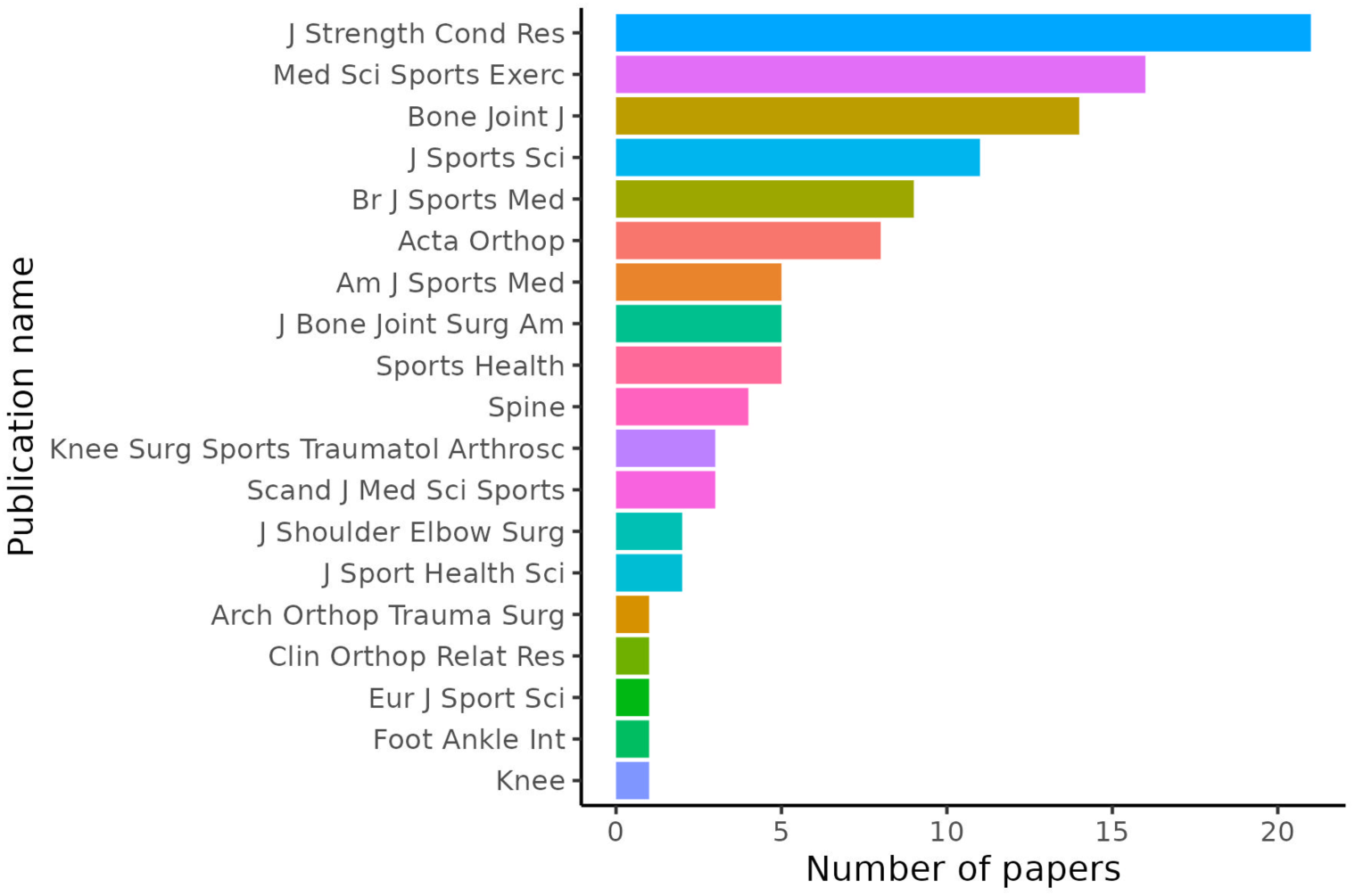
Breakdown of included papers by journal name.

**Table S1:** Description of the TRAIN and TEST datasets.

|  | **TEST,**<br>**N = 198<sup>1</sup>** | **TRAIN,**<br>**N = 784<sup>1</sup>** |
| --- | --- | --- |
| Papers (n, unique) | 96 | 113 |
| Paper Section (n, %) |  |  |
| Introduction | 23<br>(12%) | 90<br>(11%) |
| Method | 152<br>(77%) | 604<br>(77%) |
| Results | 23<br>(12%) | 90<br>(11%) |
| Questions (n, %) |  |  |
| Does the article include the study hypotheses? | 23<br>(12%) | 90<br>(11%) |
| Does the article define the primary outcome or primary endpoint? | 28<br>(21%) | 80<br>(10%) |
| Does the Method include how the sample size was determined? | 21<br>(11%) | 87<br>(11%) |
| Does the article define the study's eligibility criteria? | 22<br>(11%) | 86<br>(11%) |
| Does the article provide a detailed description of the specific techniques used for the practical implementation of the randomization sequence? | 16<br>(8%) | 92<br>(12%) |
| Does the article include the randomization methods used in the study? | 17<br>(9%) | 91<br>(12%) |
| Does the article EXPLICITLY detail the individuals or bodies or roles involved in either GENERATING the random allocation sequence, or ENROLLING participants, or ASSIGNING participants to trial groups? | 22<br>(11%) | 86<br>(11%) |
| Does the article include details about who, if anyone, was blinded to the assignment to interventions? | 26<br>(13%) | 82<br>(10%) |
| For the PRIMARY outcome, do the results reported in the article include the standardized effect size and confidence interval? | 23<br>(12%) | 90<br>(11%) |

<sup>1</sup> = n question-text pairs

**Table S2.** Full user prompt for each text reporting guideline item.

| Guideline Item | User prompt |
| --- | --- |
| The study hypotheses | *"Task: Analyze the article focusing specifically on the details of the hypotheses used in the study. Note that acceptable hypotheses are clear, testable propositions or predictions that are specifically formulated for statistical analysis. Your task is to identify whether the hypotheses are clearly stated and if they meet these criteria. If the text only mentions hypotheses in general without clearly stating them or if they are not formulated for statistical analysis, your answer should be 'NO'. Question: Does the article include the study hypotheses?"* |
| The primary outcome | *"Task: The primary outcome or primary endpoint is the pre-specified outcome considered to be of greatest importance to relevant stakeholders and is usually used in the sample size calculation. Question: Does the article define the primary outcome or primary endpoint?"* |
| The sample size | *"Task: Analyze the article focusing specifically on the details of how the sample size was determined. Question: Does the Method include how the sample size was determined?"* |
| The eligibility criteria | *"Task: Analyze the article for detailed information about the predefined study inclusion and exclusion criteria. Inclusion and exclusion criteria are specific conditions predefined by researchers that determine who can or cannot participate in the study. If the terms 'inclusion criteria' or 'exclusion criteria' are not present in the article, you must answer NO. Question: Does the article define the study's eligibility criteria?"* |
| The implementation of the randomization sequence | *"Task: Identify whether the article provides a detailed description of the specific techniques used for the implementation of the randomization sequence. Randomization techniques are practical techniques used to generate randomization sequences or store each person's random assignment. Examples include random number tables or lists, computer software or website-generated randomization methods, sealed envelopes, and coin toss. Question: Does the article provide a detailed description of the specific techniques used for the practical implementation of the randomization sequence?"* |
| The randomization methods | *"Task: Analyze the article focusing specifically on the details of the randomization method used in the study. Note that acceptable randomization methods include covariate adaptive randomization, stratified randomization, block randomization, simple randomization, minimization, factorial randomization, and cluster randomization. Your task is to identify whether any of these specific randomization methods are mentioned or described in the text. If the text only mentions randomization in general without specifying any of these methods, your answer should be NO. Question: Does the article include the randomization methods used in the study?"* |
| The random allocation sequence & assignment | *"Task: Examine the article to determine if it explicitly details the individuals or bodies or roles involved in either generating the random allocation sequence, enrolling participants, or assigning participants to trial groups in the study. Question: Does the article EXPLICITLY detail the individuals or bodies or roles involved in either GENERATING the random allocation sequence, or ENROLLING participants, or ASSIGNING participants to trial groups?"* |
| Blinding | *"Task: Analyze the article to determine if it includes details about who was blinded after the assignment to interventions in the trial. The term 'blinding' or 'masking' refers to withholding information about the assigned interventions from people involved in the trial who may potentially be influenced by this knowledge. Examples include no blinding, or participants, care providers, or assessors being blinded. If the article does not include these details, your answer should be 'NO.' Question: Does the article include details about who, if anyone, was blinded to the assignment to interventions?"* |
| Standardized effect sizes and confidence intervals | *"Task: Identify the primary outcome and determine if the article reports a standardized effect size and confidence interval for the PRIMARY outcome only. Examples include odds ratios, hazard ratios, risk ratios, risk difference, standardized mean differences, Cohen's d. Question: For the PRIMARY outcome, do the results reported in the article include the standardized effect size and confidence interval?"* |
| The number of participants randomly assigned | *"Question 1: Does this image contain a CONSORT flow diagram showing, for each group, the number of participants who were randomly assigned, received the intended treatment, and were analysed for the primary outcome?"* |
| The number of participants lost to follow-up | *"Question 2: Does this image display a CONSORT flow diagram that details both the number of participants lost to follow-up and excluded after randomization AND specifies the reasons for these losses in each group?"* |

**Table S3:**
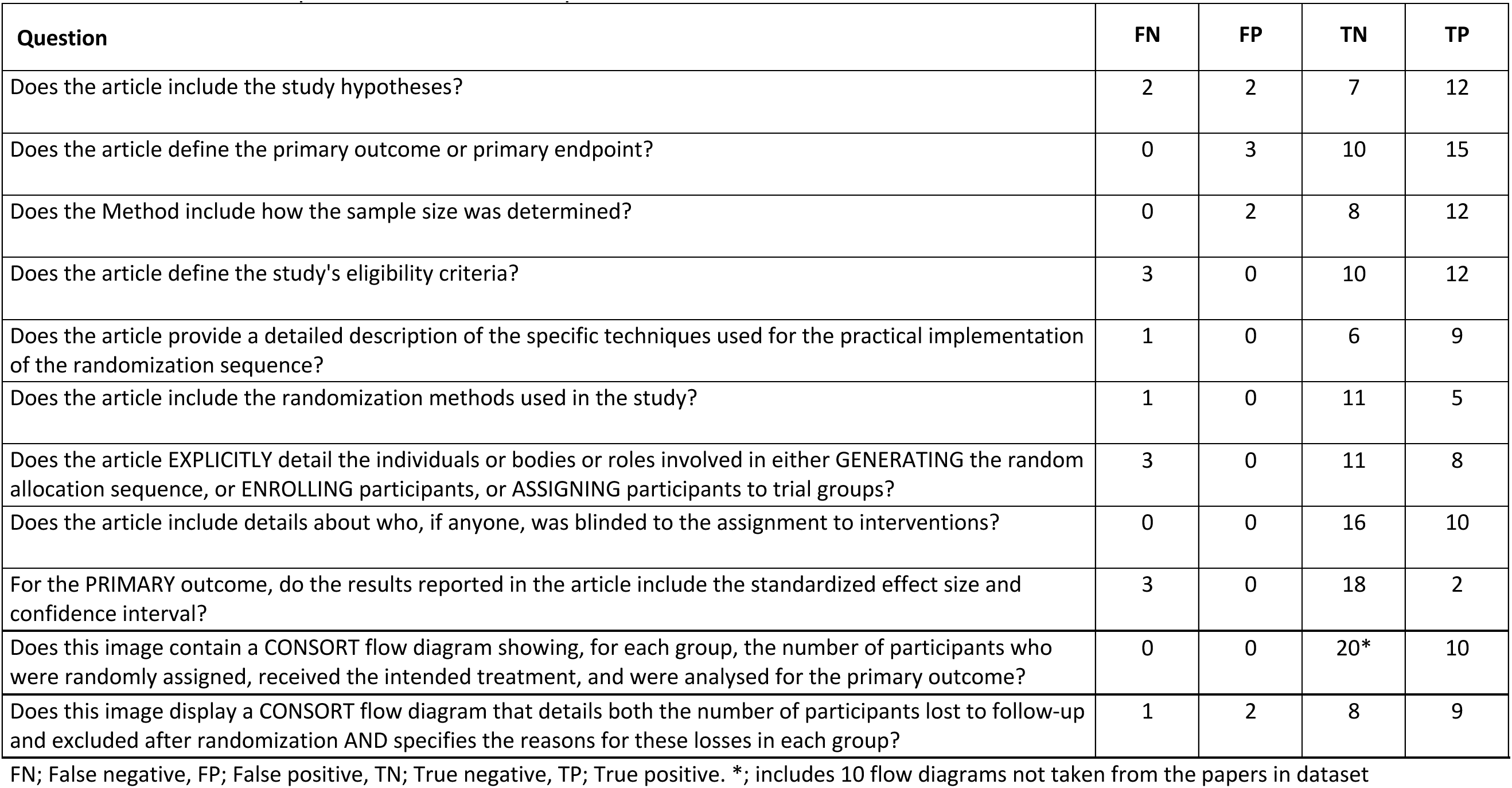
GPT-4 models performance for each question in the TEST dataset.

| Question | FN | FP | TN | TP |
| --- | --- | --- | --- | --- |
| Does the article include the study hypotheses? | 2 | 2 | 7 | 12 |
| Does the article define the primary outcome or primary endpoint? | 0 | 3 | 10 | 15 |
| Does the Method include how the sample size was determined? | 0 | 2 | 8 | 12 |
| Does the article define the study's eligibility criteria? | 3 | 0 | 10 | 12 |
| Does the article provide a detailed description of the specific techniques used for the practical implementation of the randomization sequence? | 1 | 0 | 6 | 9 |
| Does the article include the randomization methods used in the study? | 1 | 0 | 11 | 5 |
| Does the article EXPLICITLY detail the individuals or bodies or roles involved in either GENERATING the random allocation sequence, or ENROLLING participants, or ASSIGNING participants to trial groups? | 3 | 0 | 11 | 8 |
| Does the article include details about who, if anyone, was blinded to the assignment to interventions? | 0 | 0 | 16 | 10 |
| For the PRIMARY outcome, do the results reported in the article include the standardized effect size and confidence interval? | 3 | 0 | 18 | 2 |
| Does this image contain a CONSORT flow diagram showing, for each group, the number of participants who were randomly assigned, received the intended treatment, and were analysed for the primary outcome? | 0 | 0 | 20* | 10 |
| Does this image display a CONSORT flow diagram that details both the number of participants lost to follow-up and excluded after randomization AND specifies the reasons for these losses in each group? | 1 | 2 | 8 | 9 |

FN; False negative, FP; False positive, TN; True negative, TP; True positive. \*; includes 10 flow diagrams not taken from the papers in dataset

**Table S4.** Llama 2 performance for the assesed reporting guideline items.

| # | Question | Accuracy [95%CI] |
| --- | --- | --- |
| 1 | Does the article include the study hypotheses? | 83% [63-93%] |
| 2 | Does the article define the primary outcome or primary endpoint? | 93% [77-98%] |
| 3 | Does the Method include how the sample size was determined? | 86% [65-95%] |
| 4 | Does the article define the study's eligibility criteria? | 68% [47-84%] |
| 5 | Does the article provide a detailed description of the specific techniques used for the practical implementation of the randomization sequence? | 87% [62-96%] |
| 6 | Does the article include the randomization methods used in the study? | 71% [47-87%] |
| 7 | Does the article EXPLICITLY detail the individuals or bodies or roles involved in either GENERATING the random allocation sequence, or ENROLLING participants, or ASSIGNING participants to trial groups? | 76% [55-89%] |
| 8 | Does the article include details about who, if anyone, was blinded to the assignment to interventions? | 96% [80-99%] |

